# The Relationship Between Treatment Center Services and Number of Opioid-related Deaths in the United States Before and After a Declaration of a National Opioid Crisis

**DOI:** 10.1101/2022.10.03.22280663

**Authors:** Courtney L. Hatton, Brittany N. Davis, Mahamed A. Jama, Nidha S. Samdani

**Author notes:** Authors contributed equally.

## Abstract

Opioid-related deaths are a national problem that have increased over the past two decades. Multiple policy interventions have been enacted to decrease opioid misuse and expand treatment. The Comprehensive Addiction and Recovery Act (CARA) was passed in July 2016, just before declaring the opioid epidemic a National Emergency in 2017. CARA was enacted to combat the opioid epidemic by providing more funding yearly for items including but not limited to prevention, treatment, and opioid overdose reversal. To evaluate the impact of these policy changes, we carried out secondary data analysis for the period 2011-2019 using the CDC’s Wide-ranging Online Data for Epidemiologic Research and National Survey of Substance Abuse Treatment Services databases.

Research variables included: a comparison of the 50 states across the 2011-2019 timeframe, the number of opioid treatment centers, the percentage of government funding for facilities per state, percentage of opioid treatment facilities which offer free/low-income services and the opioid death rate. We also assessed differences in low-income access to opioid treatment services by comparing Medicaid expansion states versus non-Medicaid expansion states.

While both the number of treatment facilities per state and opioid death rates nearly doubled during this time, there was little to no association between them (R^2^ ranging from: 0.094-0.188 for years 2013-2019). Our research suggests that while state-level differences in opioid use disorder treatment facility characteristics related to access to care, they were only weakly associated with opioid-related deaths. This analysis may be used in the planning of subsequent actions against the national opioid epidemic and invites further inquiry into the impact of state Medicaid expansion on drug-specific opioid usage and mortality.

## INTRODUCTION

The rising opioid epidemic has become a significant public health crisis over the last decade and a half. This trend has been observed with a peak in opioid prescription dispensing in 2011, a substantial increase in opioid-related deaths since 2000, and an increase in the point prevalence of opioid use disorder (OUD) (1).

There have been multiple public policy interventions to address the epidemic at the state and federal levels. These include restrictions on prescribing opioids, along with law enforcement crackdowns of negligent prescribing (2-3). Additionally, there has been the widespread implementation of prescription drug monitoring programs (PDMPs) in 48 states by 2014, increasing from 11 in 2007 (4). A recent retrospective study on this topic found an association between state implementation of PDMPs and decreased opioid-related death rates from 1999 to 2013. The strength of this effect was moderated by drug program characteristics (4).

Other policy changes have targeted treatment and funding. The passage and implementation of the Affordable Care Act in the United States (U.S.) in 2010 has been a driving force in expanding access and treatment for substance use disorder (SUD) and OUD (5). It expanded insurance coverage to millions of Americans through Medicaid, established parity in requirements for SUD treatment for patients covered under Medicaid, reduced preauthorization requirements for OUD treatment, and added coverage for initial screenings of SUD (5-7). In addition, state Medicaid expansion has theoretically increased access to OUD treatment. However, the adoption of Medicaid expansion has been staggered and incomplete (8). In addition, the opioid overdose epidemic was declared a U.S. public health emergency in 2017, just after the Comprehensive Addiction and Recovery Act (CARA) was signed into law in 2016, both of which provided further funding for OUD treatment, prevention, and opioid-overdose reversal (9). The opioid epidemic has disproportionately affected people living in poverty and created an economic crisis for people with OUD (9).

There has been limited research on how increased funding from CARA and Medicaid expansion for OUD treatment facilities and their services have affected the opioid death rate at the state level. Nevertheless, once a person is addicted to opioids, treatment facilities are the primary option for reducing OUD and preventing opioid-related deaths (8). Combating the increase in OUD prevalence is multifaceted and warrants interventions at both the prescriber and user-level (9).

To investigate the state-level impact of CARA on OUD treatment facilities and opioid-related deaths, we analyzed publicly available data from the Centers for Disease Control’s (CDC) Wide-ranging Online Data for Epidemiologic Research (WONDER) and National Survey of Substance Abuse Treatment Services (N-SSATS) databases from 2011-2019 for all fifty states (10-11). We compared differences in opioid-related death rates based on state Medicaid expansion status and other low-income access markers such as the percentage of opioid treatment centers receiving government funding and the portion of facilities per state offering free services or accepting Medicaid payment.

Hypothesis one was that increased funding in the form of services offered by treatment facilities was associated with a decreased opioid-related death rate at the state level. Hypothesis two was that those states with a higher rate of facilities providing low-income access would have decreased opioid-related deaths compared to states with less access.

## METHODS

### Procedures

We used data from the CDC WONDER database and the National Survey Substance Abuse Treatment Services (N-SSATS) (10-11). The CDC WONDER Database data set is collected and updated yearly; the most recent data available currently is from 2019. The N-SSATS data set is compiled by the Substance Abuse and Mental Health Services Administration (SAMHSA) from a voluntary annual census of substance treatment centers throughout the U.S. It includes facility-specific responses to treatment utilization, funding types, type and the number of services offered, and other facility characteristics. We aggregated facility data to the state level. We used data from 2011 through 2019 in a zero-correlation study design with archival data. We chose this approach to determine a relationship between increased funding of substance treatment facilities and opioid-related deaths. The N-SSATS database was used to identify treatment facilities that provided OUD treatment services specifically. The percentage of those facilities in each state received government funding (state, federal or local) and which percentage offered free services or accepted Medicaid. We compared this state-level data to the CDC WONDER database to determine if there is a correlation with opioid-related deaths per 100,000 inhabitants.

Opioid-related deaths and the number of opioid treatment facilities were adjusted to reflect a per capita number. This enabled us to compare the numbers corresponding to each state and control differences in state population size. The N-SSATS survey had a variable non-response rate from facilities from year to year (Table 1) (11). For our purposes, missing data were omitted. We believe this study should be relatively resistant to outlier data because, as seen above, the missing data accounts for ∼8% of the data. Typically research with secondary data can have up to 10% missing data without affecting it statistically (12). We chose and adjusted variables to reduce confounding variables within secondary data-analysis limitations.

**Table 1.** N-SSATS Facility Respondents. Values displayed represent the total number of eligible facilities within the N-SSATS database from 2011 to 2019, including the total number of respondents, non-respondents, and percentage of non-respondents. The average percent of non-respondents is 7.73%.

FIGURES
| Year | Total Eligible Facilities | # of Non-respondents | % of Non-respondents | # of Respondents |
| --- | --- | --- | --- | --- |
| 2011 | 15,222 | 920 | 6.0% | 14,302 |
| 2012 | 16,114 | 1,119 | 6.9% | 14,995 |
| 2013 | 15,496 | 866 | 5.6% | 14,630 |
| 2014 | 15,351 | 969 | 6.3% | 14,382 |
| 2015 | 15,537 | 1,303 | 8.4% | 14,234 |
| 2016 | 16,007 | 1,375 | 8.6% | 14,632 |
| 2017 | 15,528 | 1,671 | 10.8% | 13,857 |
| 2018 | 16,365 | 1,377 | 8.4% | 14,988 |
| 2019 | 17,808 | 1,533 | 8.6% | 16,275 |

For determining if a facility is an opioid treatment center, we counted all facilities that provided one or more of the following opioid addiction treatment medications: methadone, naltrexone including extended-release, buprenorphine with or without naloxone, buprenorphine implants or extended-release pharmaceuticals, lofexidine, and/or clonidine. For determining opioid deaths, ICD10 codes X40-44, X60-64, X85, Y10-14 were used. This included all intentional suicides, unintentional overdoses, undetermined overdoses, and homicide via overdose. The categories were further sub-coded into T40 codes; T40.1 identifies overdoses from heroin. Prescription opioid deaths were designated by T40.2-4 and T40.6; these represent other opioids, methadone, other synthetic narcotics, and other and unspecified narcotics. Because opioid addiction can be to prescription opiates or heroin, we combined the totals, except where explicitly stated otherwise. The N-SSATS survey includes a question that asks whether the facility receives funding from any federal, state or local government; the answers were either yes or no. The total number of facilities answering yes to this question was used to determine the percentage of facilities in each state that receive government funding. However, the dollar amounts could not be determined. Similarly, the survey asks if the facility provides all services for free and if they accept Medicaid as payment. The number of facilities answering yes to either of these questions was totaled to calculate the percentage of facilities in each state that provide free services.

### Data Analysis

A correlation was used to examine the association between the number of opioid-related deaths to the number of opioid treatment facilities, the percentage of facilities in each state that receives government funding, and the percentage of facilities in each state that provide free services or accept Medicaid. In addition, we used multiple unpaired *t*-tests to assess significant differences between death rates in Medicaid expanded states vs. non-expanded states. We generated all statistics and figures in GraphPad Prism version 9.1.0 (216) for macOS (13), Microsoft Excel version 16.50(21061301) for macOS (14), and JMP version 16.0.0 (512340) for macOS (15).

## RESULTS

Between 2011 and 2019, the number of opioid treatment facilities doubled (4,280 to 8,437 facilities), and the number of national opioid deaths nearly tripled (23,617 to 64,404 deaths). State-specific trends in these metrics were visualized by heatmap distributions (Figures 1 and 2). Qualitatively, the increase in opioid overdose deaths was most significant in Northeastern and Appalachian states (Figure 2), while the most significant increase in treatment facilities was in the Southwest U.S. Changes in the mean distribution from year to year were substantial in both the opioid deaths and the number of facilities (p-values = <0.0001), showing an increasing number nearly every year except for 2018, being slightly lower than 2017. Comparison of the amount of money allocated to SAMHSA toward substance abuse treatment and prevention (Table 2) during this timeframe and the opioid deaths depicts a similar increasing trend (Figure 3) (16-26).

**Table 2.** Government spending on substance abuse treatment and prevention as reported by the SAMHSA from 2011 to 2019 (16-26). The values represent the total amount spent per year, the total amount allocated for treatment, the total amount allocated for prevention, and the total amount allocated for opioid-specific treatment and prevention. The total amount of government spending per year increased nearly two-fold by 2019, with funds spent on opioid-related resources. Dollar amounts are in thousands.

| **Year** | **Total Money Spent** | **Prevention** | **Treatment** | **Opioid specific** |
| --- | --- | --- | --- | --- |
| 2011 | \$2,400,414 | \$527,063 | \$1,873,351 | N/A |
| 2012 | \$2,414,914 | \$185,885 | \$2,229,029 | N/A |
| 2013 | \$2,289,905 | \$175,513 | \$2,114,392 | N/A |
| 2014 | \$2,351,270 | \$175,129 | \$2,176,141 | N/A |
| 2015 | \$2,356,467 | \$175,148 | \$2,181,319 | N/A |
| 2016 | \$2,406,643 | \$211,219 | \$2,195,424 | N/A |
| 2017 | \$2,930,375 | \$221,869 | \$2,708,506 | \$500,000 |
| 2018 | \$4,005,389 | \$248,219 | \$3,757,170 | \$1,500,000 |
| 2019 | \$4,022,079 | \$205,469 | \$3,816,610 | \$1,500,000 |

**Figure 1.**
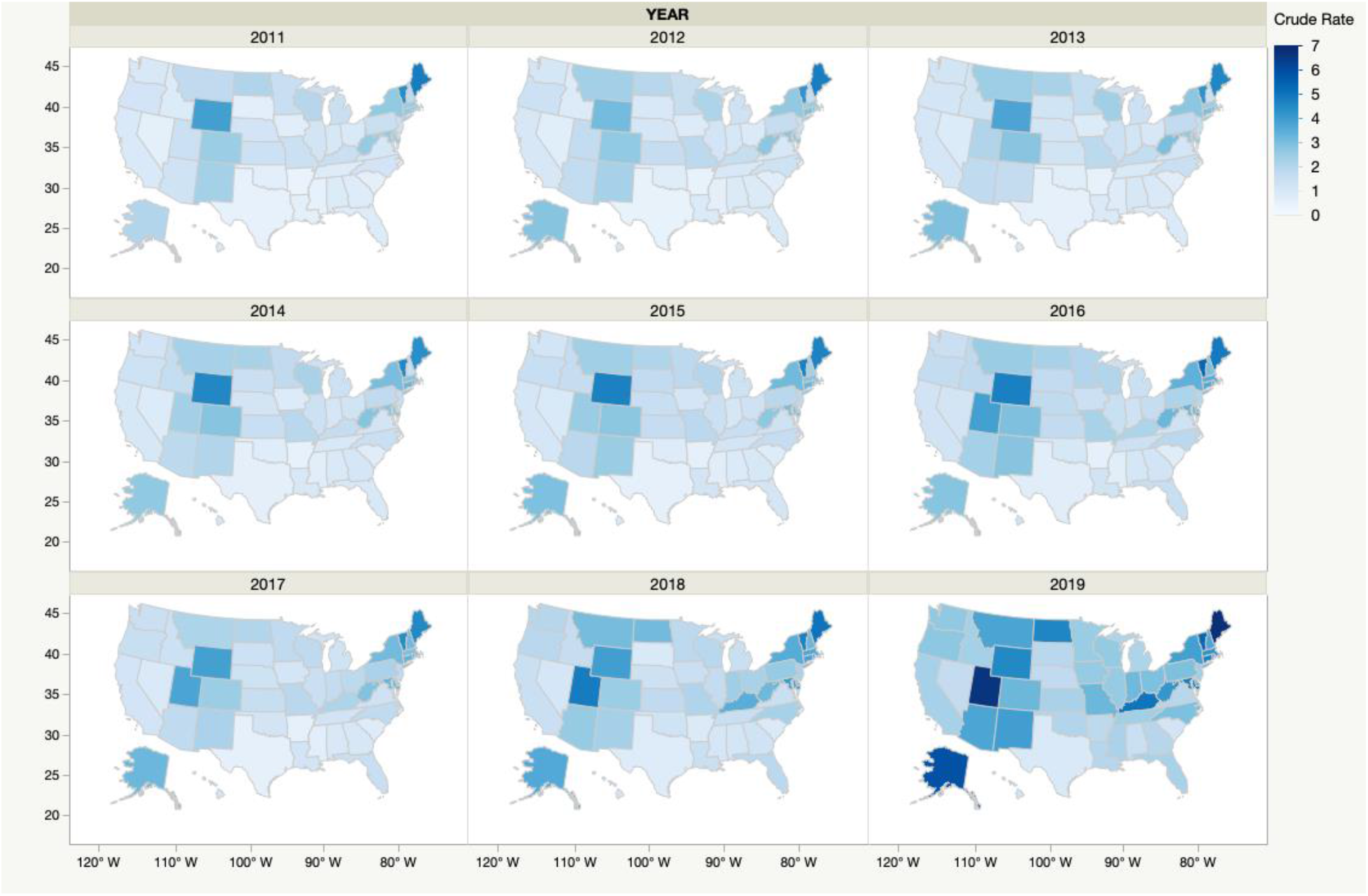
State-level heatmap of the OUD treatment facilities per 100,000 people from 2011-2019. The total number of OUD treatment facilities increased two-fold from 4,280 to 8,437.

**Figure 2.**
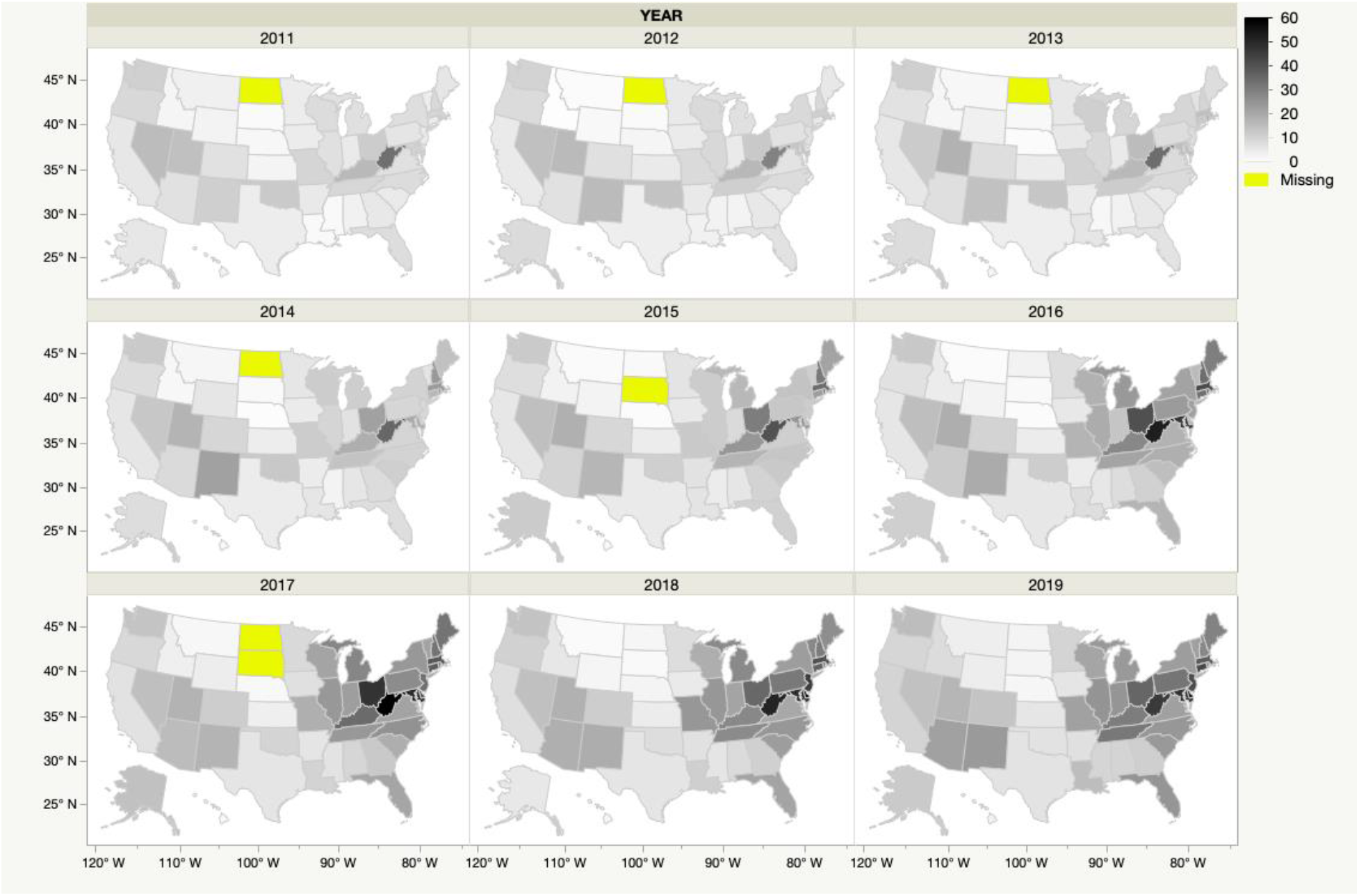
State-level heatmap of the opioid overdose deaths per 100,000 people from 2011-2019. The total number of opioid overdose deaths increased nearly three-fold from 23,617 to 64,404.

**Figure 3.**
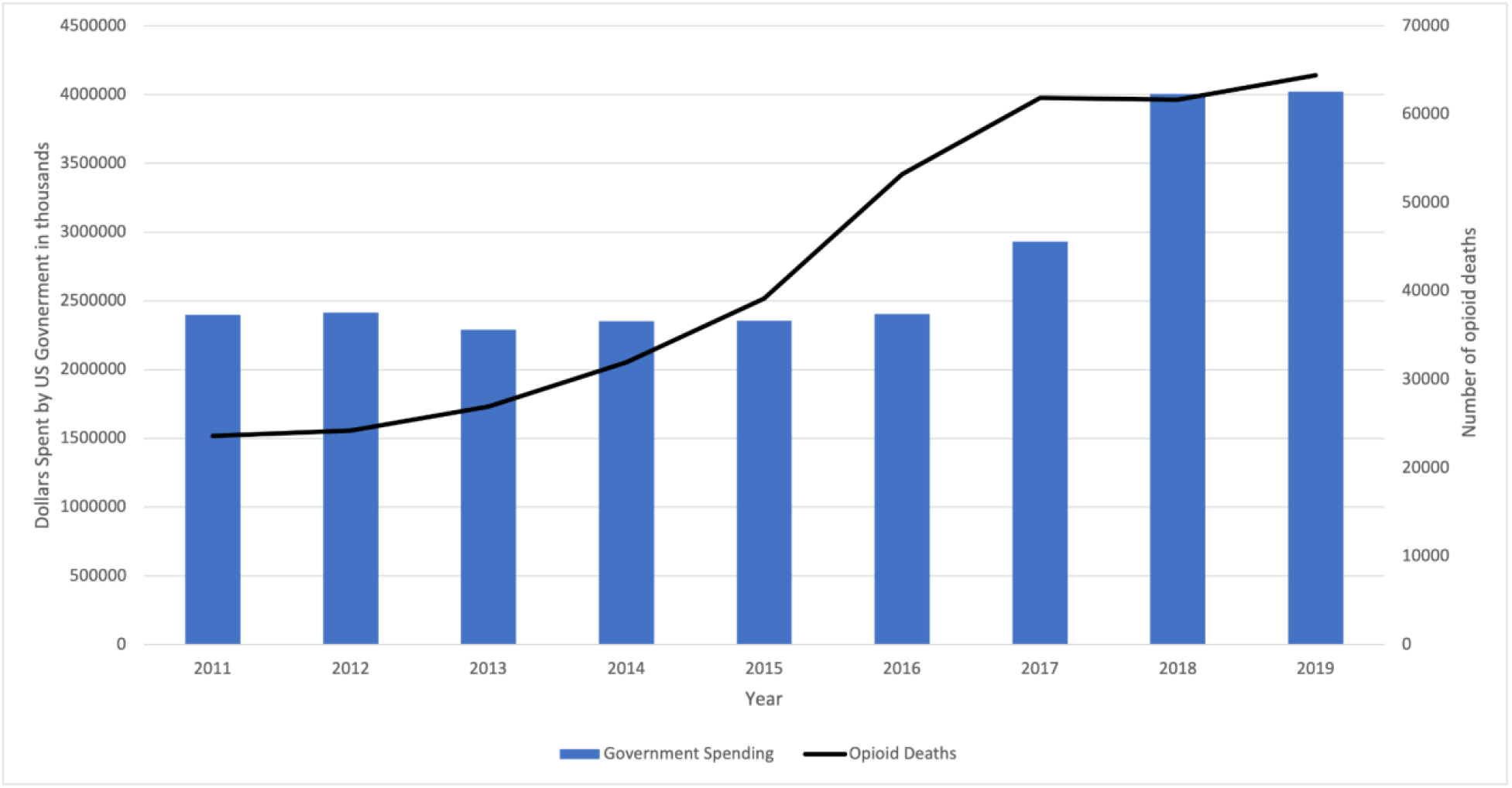
Comparison of SAMHSA allocated funding on substance abuse services and treatment versus opioid overdose deaths between 2011 and 2019. During this time, both the total amount of allocated funds for substance abuse services and opioid overdose deaths increased.

Linear regression of per capita opioid treatment centers and opioid deaths showed no to very little correlation with R^2^ values ranging from 0.018-0.192 in each year (Figure 4), and the data model was significant from 2013-2019.

**Figure 4.**
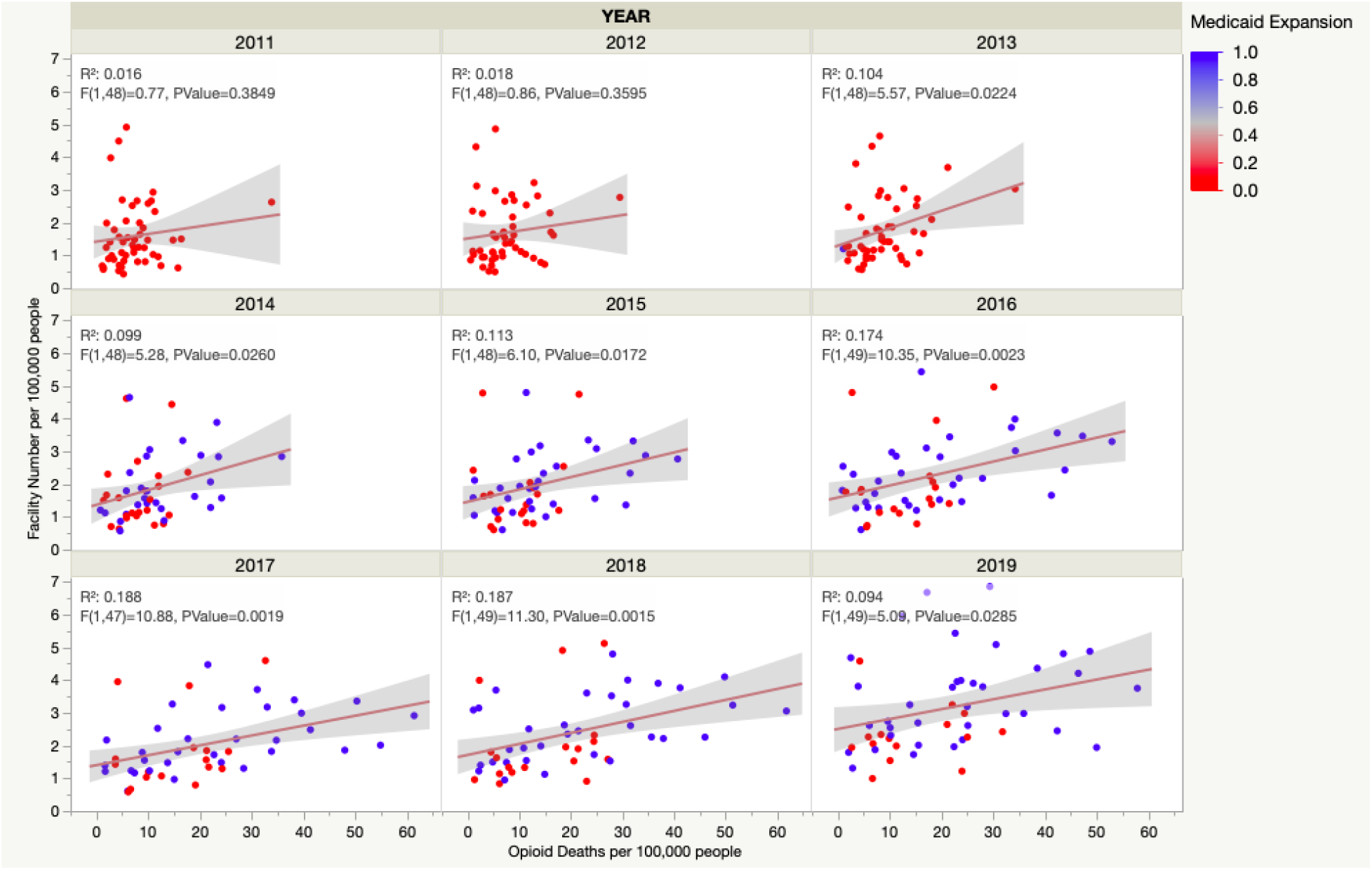
Correlation of facility crude rate numbers and opioid overdose death crude rates. Medicaid expansion was added for visualization. Linear regression of per capita opioid treatment centers and opioid deaths with R^2^ values ranging from 0.018-0.192 each year.

Other variables that could have relevance in reducing opioid deaths, such as the percentage of facilities in each state that receive government funding either from the federal, state, or local government, or the percentage of facilities in each state that provide free care to all or accept Medicaid, were examined. Each state’s range of facilities receiving government funding has remained consistent through the years, ∼25-80%, respectively. The range of facilities that provide free care or accept Medicaid has increased from ∼7-90% to ∼30-90%. This suggests that more facilities provide greater access to their services, but neither of these variables had strong correlations with the opioid death rates (Figures 5 and 6). There is no or very low correlation between opioid overdose death rates and the percentage of facilities receiving government funding or the percentage of facilities providing free services or accepting Medicaid. The government funding data was not significant, but the percentage of facilities providing free care or accepting Medicaid was significant in every year except 2011 and 2012 (P-values: 2011=0.3561, 2012=0.2464, 2013=0.0015, 2014=0.0037, 2015=0.0028, 2016=0.0101, 2017=0.0016, 2018=0.0040, and 2019=0.0150).

**Figure 5.**
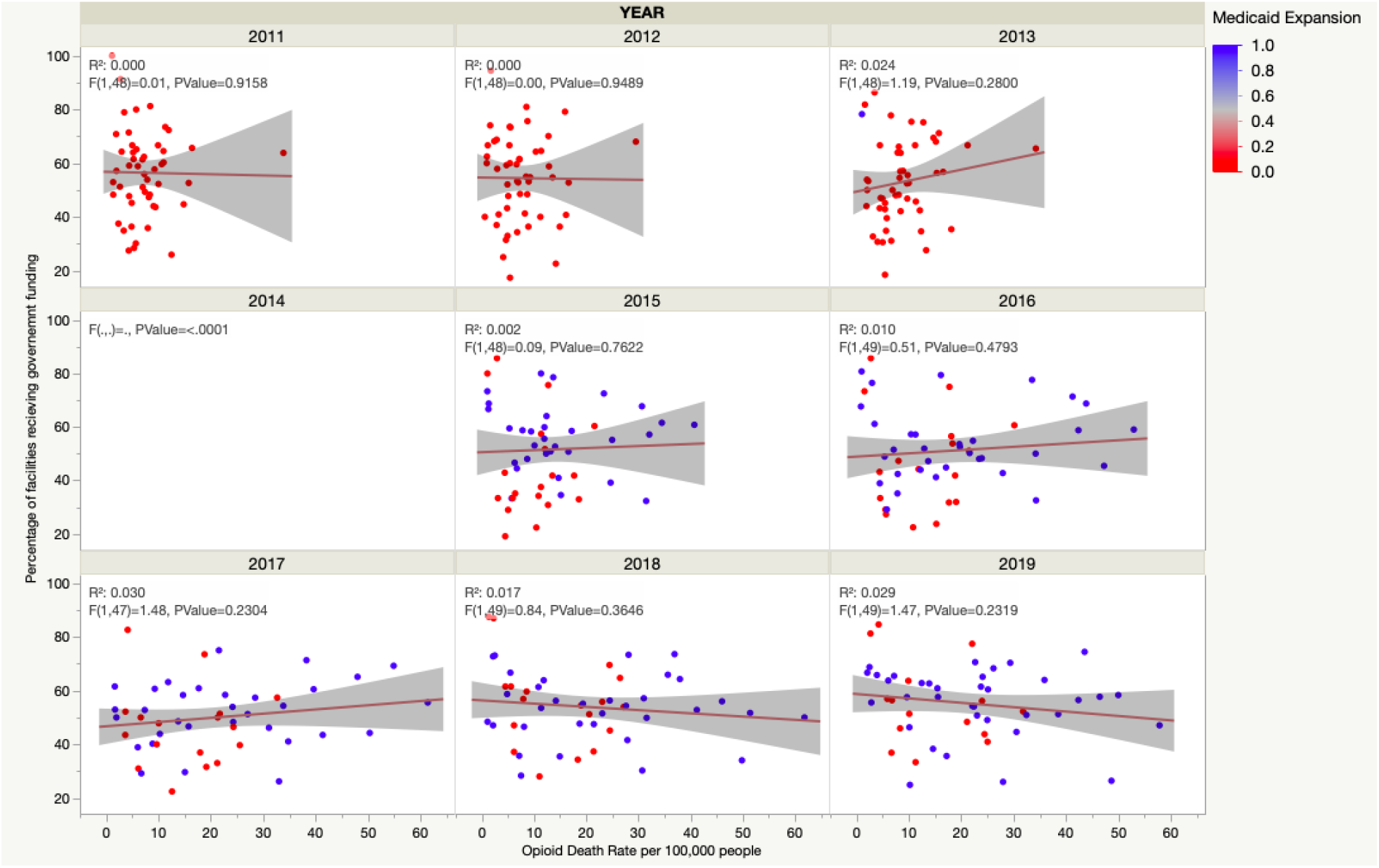
Linear regression comparisons by year for opioid deaths per 100,000 people and percent of facilities in each state receiving government funding. Data for 2014 was unavailable. There is no or very low correlation between opioid overdose death rates and the percentage of facilities receiving government funding (R^2^ values <0.03 and p-values>0.2).

**Figure 6.**
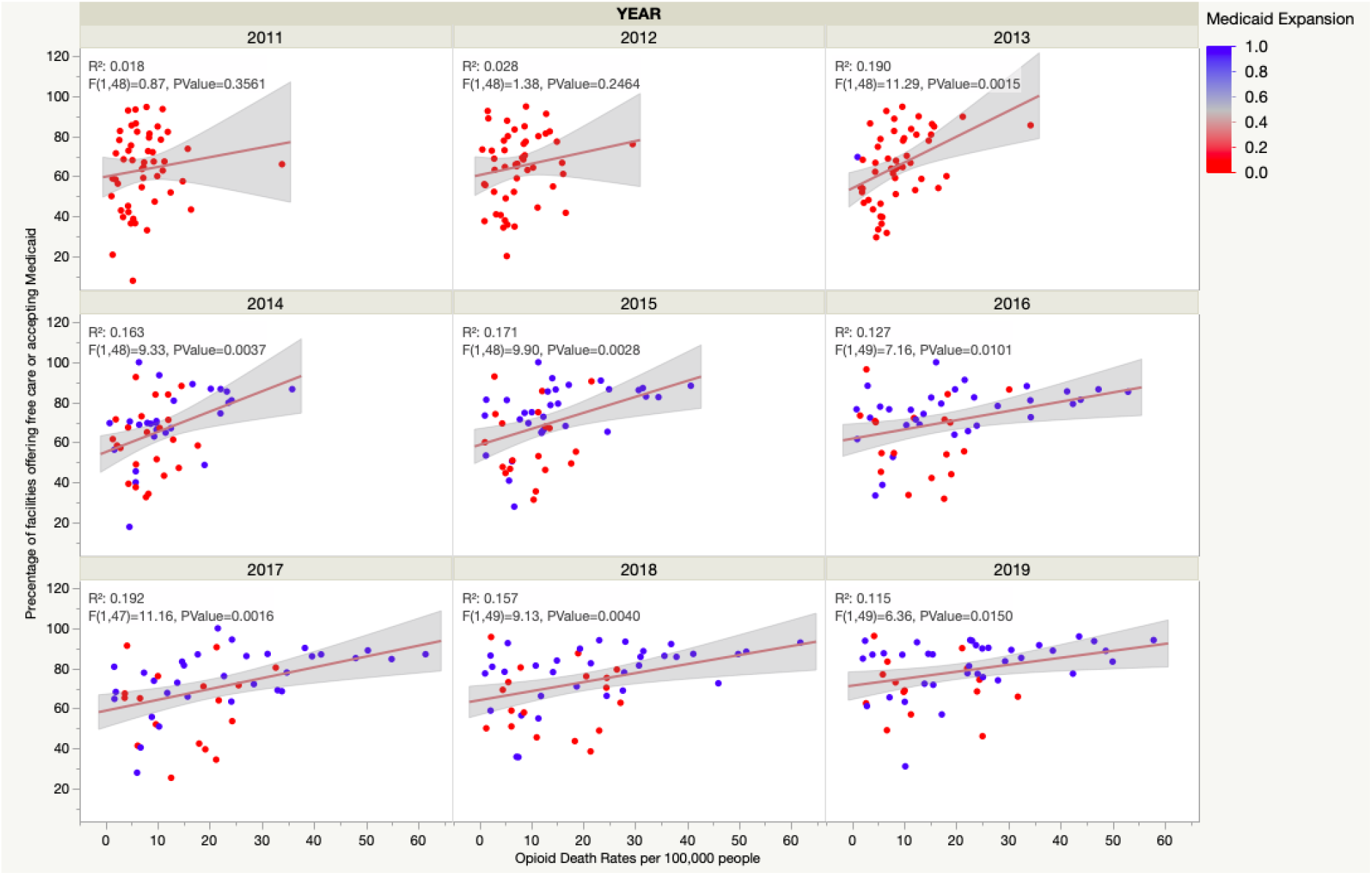
Linear regression comparisons by year for opioid deaths per 100,000 people and the percent of facilities in each state providing free or accepting Medicaid services. The range of facilities that provide free care or accept Medicaid has increased from ∼7-90% to ∼30-90%

The other factor that we hypothesized would affect the opioid overdose death rates was whether a state had expanded Medicaid or not. A comparison of the opioid death rates was made between non-Medicaid expanded states and expanded states. We determined a state was considered expanded if the expanded Medicaid plan took effect at any point in the year. Using publicly available data, we created lists of expanded states for each year (27). After comparing opioid overdose death rates between Medicaid expanded states and non-expanded states, we determined that there was no significant mean difference between the two groups for any of the years (unpaired t-test, p-value>0.05) (Figure 7A). Medicaid expansion comparisons used in Figures 4,5, and 6 were not statistically significant but were added for visualization.

**Figure 7.**
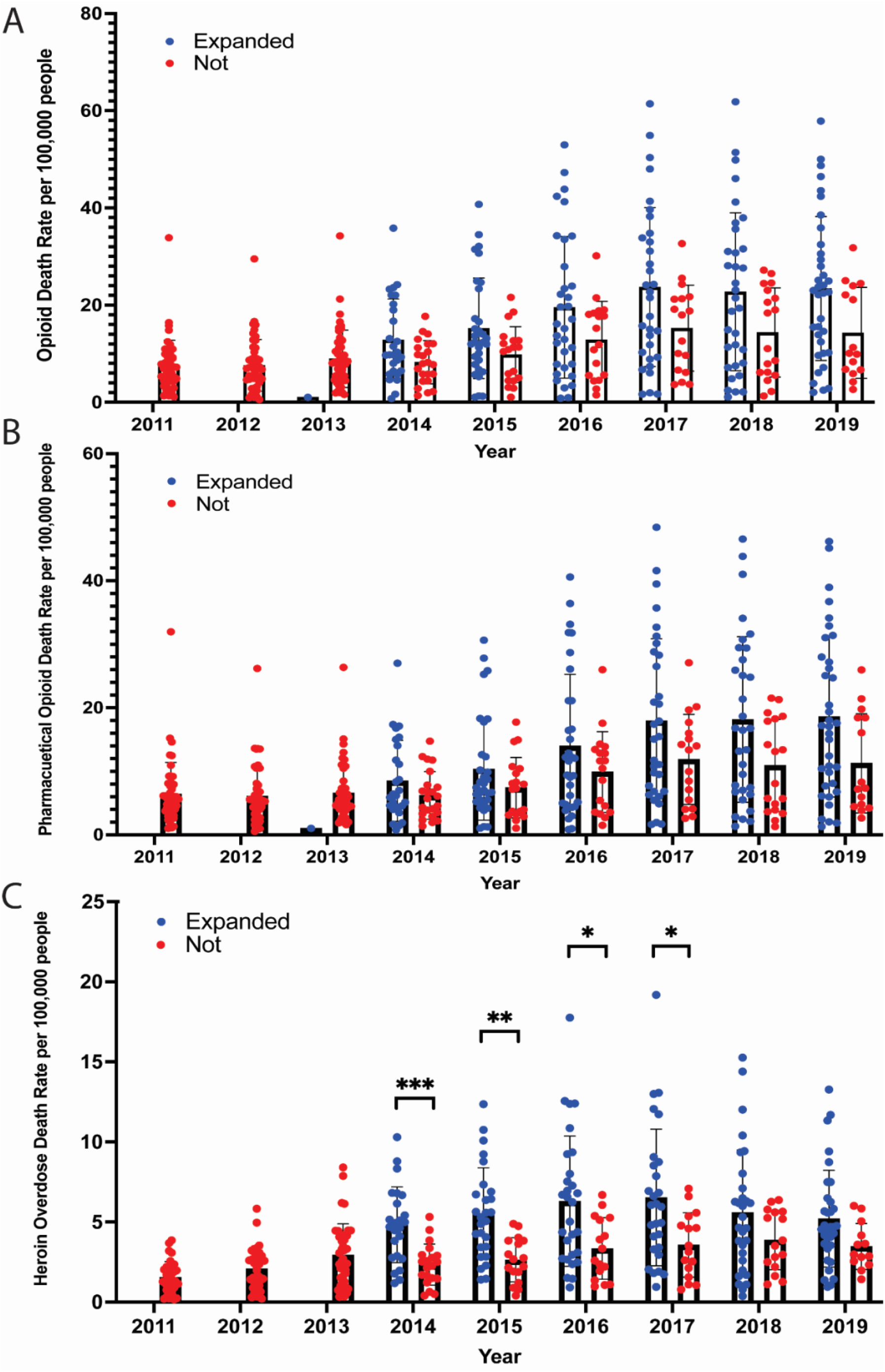
**(A)** Yearly comparison of opioid overdose deaths per 100,000 people in Medicaid expanded states versus non-expanded states. There was no significant mean difference between the two groups for any years (unpaired t-test, p-value>0.05). **(B)** Pharmaceutical opioid overdose death rates comparison between Medicaid expanded states and non-expanded states. Pharmaceutical opioid overdose deaths showed no statistically significant difference in means (unpaired t-test, p-value>0.05). **(C)** Heroin overdose death rates comparison between Medicaid expanded states and non-expanded states. The Medicaid expanded states had higher means, statistically significant in 2014, 2015, 2016, and 2017 (p-values as appears: 0.0007, 0.0017, 0.0358, and 0.0370).

We then evaluated pharmaceutical opioid overdose deaths and heroin overdose deaths separately, comparing Medicaid expanded and unexpanded states (Figures 7B and 7C). Like the total opioid overdose deaths, pharmaceutical opioid overdose deaths showed no statistically significant difference in means. However, in the heroin overdose death rates, the Medicaid expanded states had higher means, and they were statistically significant in 2014, 2015, 2016, and 2017 (p-values as appears: 0.0007, 0.0017, 0.0358, and 0.0370). In the Medicaid expanded states, though, there seemed to be a leveling of deaths after 2017. Additionally, the heroin deaths in the Medicaid expanded states seemed to be getting lower. Again, it was not statistically significant and could be due to multiple missing data points that could not be determined if it was non-reported or reported zero.

Finally, we qualitatively visualized the effect of state-level Medicaid expansion using the percent change in opioid overdose deaths per 100,000 people (Figure 8). States with Medicaid expansion appear to have decreased percent changes in recent years, suggesting that though Medicaid expanded states still have more deaths, deaths decreased in those states.

**Figure 8.**
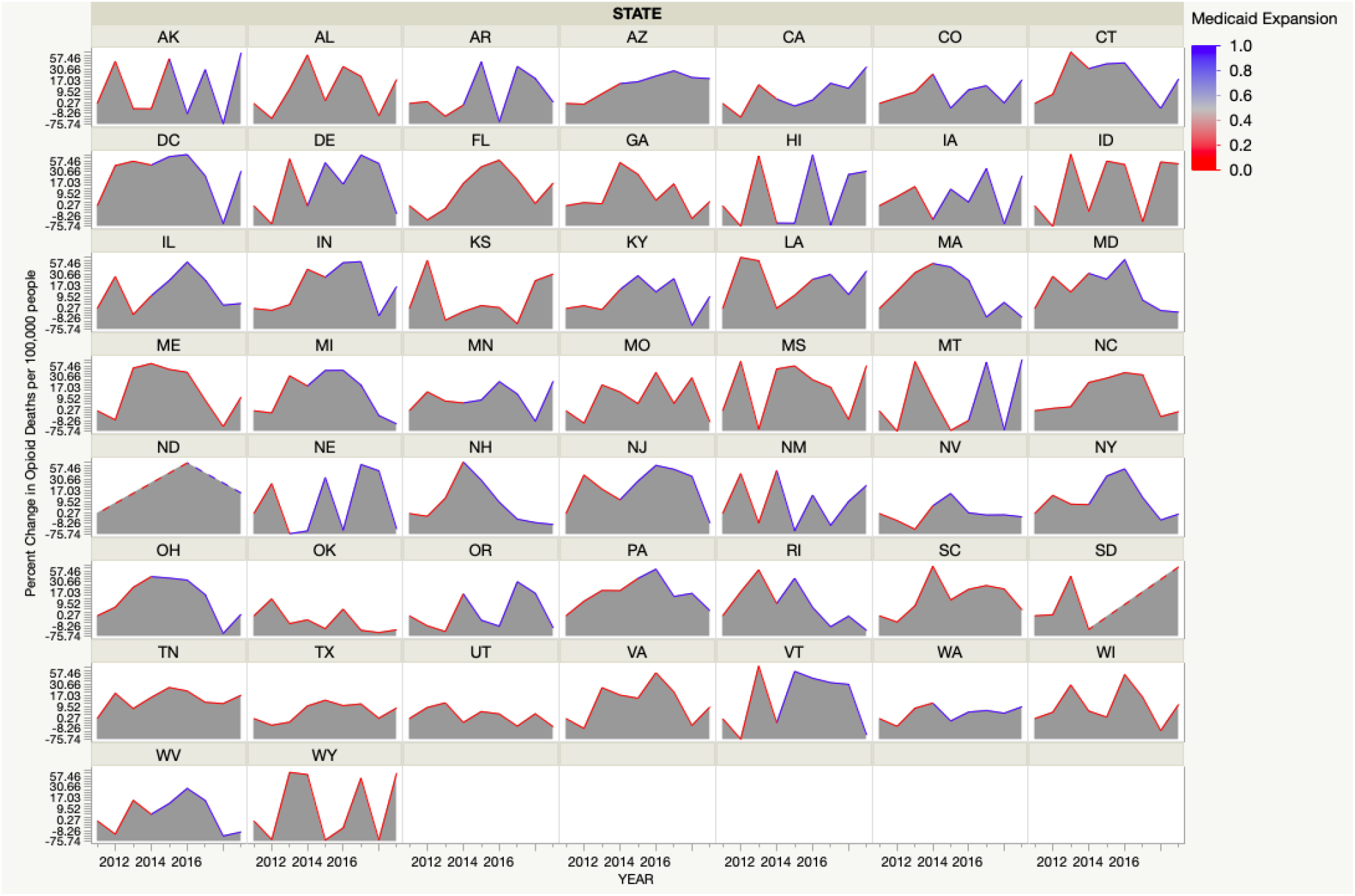
Percent change in opioid overdose death rates per 100,000 people in each state with the indication of when each state expanded Medicaid.

## DISCUSSION

The results of our research did not support our first hypothesis that a higher number of facilities providing low-income access would decrease opioid-related deaths versus states which do not. Overall, our data showed that the number of opioid-related deaths has almost tripled from 2011 to 2019. The results did not indicate any pattern between opioid-related deaths and the number of facilities, nor increased funding and opioid-related deaths. However, the results showed an increase in opioid-related deaths and an increase in the number of facilities per capita.

Although our data showed an increase in the number of treatment centers and opioid-related death rates, there seems to be a question about the efficiency of these current treatment centers. The ACA aims to address the opioid crisis through funding, ultimately leading to increasing treatment facilities (28). However, increased opioid treatment centers do not correlate to the increased treatment availability for all users. Despite the increase in facilities, only 20% of opioid users receive treatment (28). In addition, there are barriers for opioid users seeking treatment, such as being placed on waitlists for programs that can last days, up to months. This can lead to continuous unguided drug usage behavior, which can contribute to opioid-related deaths. A method needs to be implemented to optimize these treatment centers for those currently on the waiting lists to address facilities’ efficiency. There are current efforts of a technology-assisted interim dosing regimen (29). This includes creating methods to monitor patients through computerized buprenorphine dispensing and contacting patients through phone communication and random call-backs. This method has demonstrated efficacy in reducing opioid and drug use behavior and decreasing psychiatric distress during the waitlist process. Another issue is opioid users need a holistic, tailored treatment based on their history and opioid usage behavior. Vermont is one of the states that has developed a “hub-and-spoke system” which allows patients to start more intensive and custom treatment through in-person counseling, urine toxicology testing, and other medical management (29). In addition, Vermont aims to provide effective treatment by implementing a brief screening questionnaire to help match the most appropriate treatment to each patient (28). This information helps us better understand why increased treatment centers do not decrease opioid-related deaths. There needs to be an internal evaluation of current treatment facilities by addressing their waitlist patients, creating personalized treatments, and providing a screening method to match patients to the appropriate treatment type.

In addition to facility efficacy, another factor affecting the correlations between facility number and opioid-related deaths could be due to how we calculated whether a facility was an opioid treatment facility or not. We classified a facility as an opioid treatment facility if their response to the N-SSATS survey indicated that they provided one or more of the following opioid addiction treatment medications: methadone, naltrexone including extended-release, buprenorphine with or without naloxone, buprenorphine implants or extended-release pharmaceuticals, lofexidine, and/or clonidine. This was the best method of interrupting the data. Still, it likely included facilities that supply the pharmaceuticals but do not provide other necessary treatment services, such as mental and social health services. Additionally, because the N-SSATS survey is only given to physical facilities, it excludes the 37,000+ providers that SAMHSA provides waivers and certifications to, allowing them to prescribe opioid treatment medications (24). The opioid-related deaths are not declining, but perhaps if there were a better way of determining the number of patients seen in facilities and private providers, there would be a better way to determine a correlation between treatment and opioid-related deaths.

The lack of significance between Medicaid expanded and unexpanded states and the fact that expanded states have higher means could be explained by larger states having expanded sooner than smaller states. Even though the data was adjusted based on population size, states with larger populations would have a more significant load on the Medicaid system, so the effect of Medicaid expansion might be less visible initially.

Additionally, since we only looked at whether a state expanded Medicaid and not how it expanded, there are likely variables within the difference expansions that could act as confounding factors. Likewise, we did not investigate the coverage available before the expansion and what coverage is available in non-expanded states. Since each state individually decides on its Medicaid expansion coverage, it is not a uniform variable and therefore makes the evaluation of its effectiveness difficult.

An additional factor that we did not assess in this study but is a topic for future research is the use of opioid antagonists. SAMHSA has been promoting the training and use of opioid antagonists, such as naloxone, which can save a person’s life who is actively overdosing on opioids. Since these medications can make the difference between life and death, their use and implementation across the country could be a better predictor of opioid death rates. In a 2018 Morbidity and Mortality weekly report, the CDC investigated the emergency use of naloxone and found that its use rose by 75.1% from 2012-2016 (30). The authors used the emergency medical service records to collect this data. As a future addition to this study and our study, the use of naloxone should be investigated to determine its correlation with opioid-related deaths.

## CONCLUSIONS

This study evaluated multiple ways in which increased access to OUD treatment services influenced opioid-related death rates. There are state-level differences in OUD treatment facility characteristics associated with opioid-related mortality, but these are only weakly correlated with opioid-related death rates. Medicaid expansion was hypothesized to impact decreasing opioid-related deaths because it would allow for more access to treatment. Still, facility limitations are likely the bottleneck in treatment, not financial access. Further research is needed to draw definitive epidemiological conclusions on the impact of CARA and Medicaid expansion. However, the exploratory analysis carried out in this study can help inform future investigation and public policy aimed at addressing the opioid epidemic.

## Data Availability

All data produced in the present study are available upon reasonable request to the authors.

http://wonder.cdc.gov/ucd-icd10.html

https://www.datafiles.samhsa.gov/study-series/national-survey-substance-abuse-treatment-services-n-ssats-nid13519

## Acknowledgments

The authors would like to thank Brian Piper, Ph.D., MS & Reema Persad-Chem, Ph.D., MPH, for their invaluable guidance and assistance throughout the research process.

## Disclosures

The authors have no conflicts of interest to disclose.

## Notes

### Competing Interest Statement

The authors have declared no competing interest.

### Funding Statement

This study did not receive any funding.

### Author Declarations

IRB of Geisinger Commonwealth School of Medicine gave ethical approval for this work, approval #2021-0794.

